# Characterization of groups of informal caregivers. A cluster analysis using the example of Saxony

**DOI:** 10.1101/2023.11.01.23297767

**Authors:** Tim Tischendorf, Silke Geithner, Tom Schaal

## Abstract

**Background:** As a result of demographic change, a further increase in the number of people in need of care in Germany can be expected in the future. Nursing activities performed by family members are a central component of care provision. The assumption of support services is increasingly associated with an additional physical and psychological burden on informal caregivers. The aim of this study is to identify and characterize groups of informal caregivers with regard to their well-being.

**Methodology:** The empirical study was based on a survey on home care in Saxony from 2019, which was intended as exploratory work to provide insights into the care situation in Saxony. The central component is a two-step cluster analysis with exclusively informal caregivers.

**Results:** The net sample size for the cluster analysis comprised 178 subjects who were involved in caring for relatives. The cluster analysis revealed two groups of caregiving relatives in Saxony, which were differentiated by a different experience of stress and various sociodemographic factors.

**Discussion:** Informal caregivers in Saxony are not a homogeneous group. Depending on various sociodemographic factors or the care effort and situation, they are confronted with different challenges in caring for relatives, which are directly reflected in their well-being. In order to achieve a targeted reduction in the burden on family caregivers, cooperation and constructive collaboration between political decision-makers, care and health insurers, and the various players in health and care provision is required.

## Introduction

Germany is in the middle of demographic change. This is characterized by a decreasing number of young people and a simultaneous growth in the number of older people [1]. The impact of demographic change varies from region to region. In a nationwide comparison, the situation is more acute in Saxony. While the average age of the population in many German states is between 42 and 45 years, the population living in Saxony is older than average at 46.9 years [2].

One of the reasons for demographic change is the increasing life expectancy of the population as a result of advances in medical care, hygiene, etc. [3]. According to the results of the 2018 / 2020 mortality table, average life expectancy is currently 83.4 years for newborn girls and 78.6 years for newborn boys, and has more than doubled in Germany since statistical records began [3]. Increasing life expectancy does not correlate with better health [4]. It increases the probability of the occurrence of multimorbidity, i.e., simultaneous illness from two or more chronic diseases [5]. In view of the demographic development, a further increase in the number of persons with age- and disease-related limitations can be assumed in the future. Expressed in figures, this means for Saxony in 2021 a number of persons in need of care according to SGB XI of more than 310,000 people, which means an increase of 23.9% compared to 2019 [6].

Family caregivers are a central component of care provision in Germany. This group includes family members or other people close to the person in need of care. In 2021, there were 4.96 million people in need of care in Germany, of whom 4.17 million (84%) were cared for at home and 790,000 (16%) in institutions [3]. Of the persons in need of care who were cared for at home, 3.12 million were cared for primarily by relatives, without additional outpatient services [3]. The assumption of care services by relatives not only represents the preferred wish of persons in need of care, but also contributes to the maintenance of the health and social system. However, as a result of the involvement of relatives in caregiving activities, they are increasingly confronted with an additional physical as well as psychological burden, which can promote the exposure of corresponding secondary diseases [7; 8]. Supportive measures for the informal caregivers enable the maintenance of the well-being as well as the health of the lay caregiver. For this reason, it is particularly important to identify and characterize the groups of informal caregivers in more detail in order to support the ability and willingness of caregiving relatives to take over home care in a goal- and resource-oriented manner.

In Saxony, there are currently no representative survey data that provide reliable information on the well-being of informal caregivers. The specific stresses in the various caregiving situations are insufficiently taken into account. The aim of this work was to identify the different views on well-being when taking over caregiving services from the perspective of informal caregivers. On this basis, specific burdens of caring relatives in different care situations can be better understood and highly stressed groups can be characterized more closely. Health insurance funds and local authorities can use these findings to design effective support services for highly stressed groups of family caregivers, among others, in a target- and resource-oriented manner.

## Methodology

The present, empirical study was based on a survey on home care in Saxony 2019 and was intended as exploratory work to provide insights into the care situation in Saxony (population descriptive study). The survey, on which the dataset used is based, was conducted from 1^st^ June to 21^st^ December 2019 as a cross-sectional study and sample survey. The sample was drawn as active recruitment via the registration offices in the Free State of Saxony. The random selection of the study units was not done directly from the total population, but was collected as a stratified random sample according to the number of inhabitants [9].

Data were collected using a scientific questionnaire with 68 items divided into five categories, which was aimed at both informal caregivers and non-caregiver relatives. The Burden Scale for Family Caregivers (BSFC) was used as the basis for a valid assessment of well-being. As a scientific measuring instrument, the BSFC enables the subjective burden of family caregivers to be determined. In order to determine differences in the well-being of the participants, the German version, the Häusliche-Pflege-Skala (HPS), was adapted [10]. Within the questionnaire, there was a filter question, allowing differentiation between non-caring and caring relatives. The questionnaire was checked for completeness and comprehensibility in a pretest by a questionnaire conference and adapted or improved according to the weak points [9].

The questionnaire was made available in online or paper format. In compliance with valid procurement and data protection regulations, invitation cards for the survey were sent by mail to individuals from the cleaned reporting data set as part of commissioned data processing. These contained the link to the survey as well as information on data protection, anonymity and voluntariness, whereby the respondents had to agree in writing. In addition, a hotline was set up for people who could not participate in the survey online. In order to reach these persons nevertheless, the questionnaire was sent by post with a stamped return envelope after a telephone request (n=599) [9].

The data analysis was carried out with the statistical software SPSS 29 and represented a secondary analysis of a data set that was not self-collected [9]. The target group for the cluster analysis was defined as persons of age who have cared for a relative, friend or neighbor in their own home on an approximately weekly basis. A selection of variables was made for the cluster analysis, which were eligible for the procedure and the pursuit of the objective and research question. In the first step, the three reduction criteria for the selection of variables included the inclusion of only informal caregivers on the basis of the filter question in the questionnaire. Subsequently, the selection of variables was based on the scientific work on highly stressed groups of caring relatives - results of a cluster analysis by Bohnet-Joschko [11]. In order to establish comparability of the results, those variables were left in the cluster model which were identical or had a high intersection compared to the variable selection of Bohnet-Joschko. In the final step, variables were selected and excluded from further analysis if they contained at least 25 percent missing values. In sum, the final model for cluster analysis contained nine variables, with a total of 21 inputs (Table 1).

**Table 1.** Overview of the cluster model and corrected scale levels.

| Category of questionnaire | Question | Variable |
| --- | --- | --- |
| Sociodemographic background | 51 | own gender |
|  | 52 | own year of birth |
|  | 55 | own family status |
| Statements of informal carers | 7 | relationship to the person in need of care |
|  | 11 | areas of restriction of the person in need of care |
|  | 12 | duration of support for the person in need of care |
|  | 14 | weekly amount of care required |
| General statements on the topic of care | 49 | own financial situation |
|  | 50 | own well-being |

Two-step cluster analysis was used as the cluster analytical method. The noise processing for cluster analysis was set at 25 percent [12]. Since the cluster analysis was performed using both categorical and continuous variables, the log-likelihood measure was used. The maximum number of clusters was set at 15. Cluster differences for the well-being metric variable were tested for significance. For this, we first had the skewness and kurtosis output in SPSS and then performed the Shapiro-Wilk test [13]. The significance level for the multivariate analyses was set at p<.05.

## Results

All persons in the identified sample (N=24,018) were invited to participate in the survey. A total of 1,700 valid questionnaires (online and paper versions) were available for evaluation after data cleaning. The net response rate was 7.1449 percent (n=23,793) [14]. 1,297 questionnaires (76.3%) were completed online, and 403 paper questionnaires (23.7%) were returned. To assess the representativeness of the sample, the number of inhabitants in Saxony according to three age groups on December 31, 2019 was used as a reference (n=2,460,993) [15]. The sample was representative according to the number of inhabitants living in Saxony by age group and in this respect allows conclusions to be drawn about the population living in Saxony [14] (Figure 1).

**Figure 1:**
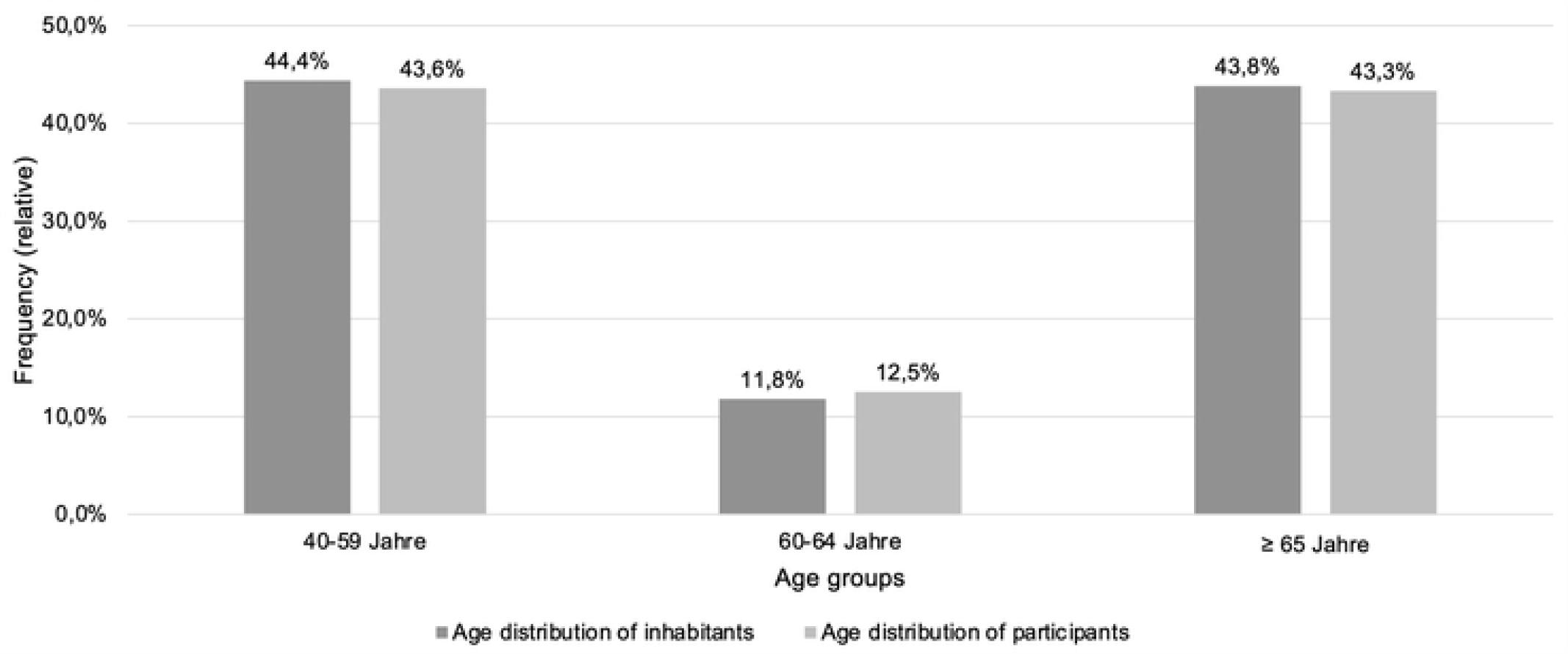
Relative frequencies of participants divided according to age groups compared with the number of inhabitants living in Saxony according to age groups (own representation).

Of the total of 310 persons who had indicated that they had cared for a relative, friend or neighbor in their home or in their own home on an approximately weekly basis in the past twelve months, 128 cases were excluded from the further investigation process because they had not answered at least one of the questions relevant to the model. The net sample size for the cluster analysis thus comprised 178 subjects.

The proportion of women was 64.0 percent (Table 2). The age of the respondents varied between 39 and 84 years (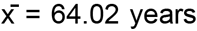 (SD ± 0.618)). 80.3 percent lived in a partnership and 5.1 percent were single. Regarding the relationship with the person in need of assistance, the majority of respondents (57.9%) reported caring for their (mother-in-law) or (father-in-law). 56.7 percent said they had been assisting the person in need of care for one to six years. In this context, 36.0 percent indicated a weekly care effort of five to less than ten hours and another 20.8 percent indicated an effort of ten to less than 20 hours. With regard to the assessment of their own financial situation, 47.8 percent rated it as “I can manage on the whole” and 29.2 percent as “I am well provided for and can afford quite a bit.”

**Table 2.** Overview of clustering characteristics.

| Characteristics | Cluster |  | Total |
| --- | --- | --- | --- |
|  | 1 (42,7%) | 2 (57,3%) |  |
| Gender |  |  |  |
| female | 59,2% | 67,6% | 64,0% |
| male | 40,8% | 30,4% | 34,8% |
| diverse | 0,0% | 2,0% | 1,1% |
| Age |  |  |  |
| 18 to 25 years | 0,0% | 0,0% | 0,0% |
| 26 to 35 years | 0,0% | 0,0% | 0,0% |
| 36 to 45 years | 5,3% | 6,9% | 6,2% |
| 46 to 55 years | 20,8% | 30,4% | 26,4% |
| 56 to 65 years | 34,1% | 44,1% | 40,0% |
| over 65 years | 39,4% | 19,0% | 27,7% |
| Family status |  |  |  |
| single | 1,3% | 7,8% | 5,1% |
| married / living in partnership | 86,6% | 75,5% | 80,3% |
| divorced / separated | 2,6% | 10,8% | 7,3% |
| widowed | 9,2% | 5,9% | 7,3% |
| Nature of the relationship |  |  |  |
| Married partner/life partner | 13,2% | 18,6% | 16,3% |
| Mother (in law) / father (in law) | 60,5% | 55,9% | 57,9% |
| (In-law / godfather / foster) child | 6,6% | 8,8% | 7,9% |
| Other relative (e.g. uncle / aunt, sister / brother, grandchild) | 6,6% | 4,9% | 5,6% |
| Friend | 1,3% | 3,9% | 2,8% |
| Neighbour | 3,9% | 1,0% | 2,2% |
| Other | 7,9% | 6,9% | 7,3% |
| Areas of limitation of the person in need of care |  |  |  |
| physical | 51,3% | 45,1% | 47,8% |
| cognitive / psychological | 7,9% | 7,8% | 7,9% |
| both | 40,8% | 47,1% | 44,4% |
| do not know | - | - | - |
| Duration |  |  |  |
| under 1 year | 21,1% | 13,7% | 16,9% |
| 1 to under 3 years | 31,6% | 22,5% | 26,4% |
| 3 to under 6 years | 30,3% | 30,4% | 30,3% |
| 6 to under 9 years | 7,9% | 11,8% | 10,1% |
| 9 years and more | 9,2% | 21,6% | 16,3% |
| There is no care level. | - | - | - |
|  | 1 (42,7%) | 2 (57,3%) |  |
| I do not know | - | - | - |
| **Weekly maintenance effort** |  |  |  |
| under 5 hours | 22,4% | 11,8% | 16,3% |
| 5 to under 10 hours | 44,7% | 29,4% | 36,0% |
| 10 to under 20 hours | 18,4% | 22,5% | 20,8% |
| 20 to under 30 hours | 9,2% | 15,7% | 12,9% |
| 30 to under 40 hours | 2,6% | 8,8% | 6,2% |
| 40 hours and more | 2,6% | 11,8% | 7,9% |
| **Financial situation** |  |  |  |
| It's not enough at all. | 1,3% | 2,0% | 1,7% |
| I just about manage to get by. | 6,6% | 27,5% | 18,5% |
| On the whole, I manage. | 32,9% | 58,8% | 47,8% |
| I am well provided for and can afford a lot. | 52,6% | 11,8% | 29,2% |
| I don't have to restrict myself in any way. | 6,6% | 0,0% | 2,8% |
| **Own well-being in the last four weeks - median figure** |  |  |  |
| I have enough time for my own interests and needs. | 3 | 2 | 3 |
| I often feel physically exhausted. | 2 | 3 | 3 |
| Every now and then I have the desire to "break out" of my situation. | 2 | 3 | 2 |
| I can be happy from the heart. | 4 | 3 | 3 |
| I sometimes no longer really feel like myself. | 1 | 3 | 2 |
| My standard of living has decreased in recent years. | 1 | 2,5 | 2 |
| I sometimes feel taken advantage of by people I support. | 1 | 2 | 1 |
| My current tasks take a lot of my own energy. | 2 | 3 | 3 |
| I feel "torn" between the different demands of my environment (e.g. work, family, care). | 2 | 3 | 3 |
| I have the feeling that I have everything "under control". | 3 | 2 | 3 |
| Because of my current tasks, my relationship with family members, relatives, friends and acquaintances suffers. | 1 | 3 | 2 |
| The fate of sick people around me makes me sad. | 3 | 3 | 3 |
| I get recognition / gratitude through my achievements. | 3 | 3 | 3 |

Note. 1 – not true; 2 – little true; 3 – mostly true; 4 – true exactly.

Information such as “[I] don’t know” or, in the case of the duration category, “There is no care level” were considered missing values in SPSS and excluded from further cluster processing.

### Groups of caregivers

The cluster analysis resulted in two groups of caring relatives in Saxony, which are shown in Table 2. The silhouette coefficient, for assessing cluster homogeneity, was in the middle range with a value of 0.3. No normal distribution could be found for the variable of well-being (p<.001). Due to the skewed distribution of values and the increased number of outliers, the median was used for the evaluation of the variable of well-being. So-called outliers can affect the average result. In contrast to the mean, these leave the median unaffected [9].

### Cluster 1: low assistance, good financial situation, high well-being

Cluster 1 was assigned 42.7 percent (n=76) of informal caregivers (Table 2). The mean age of this group was 62.12 years (SD ± 1.076). The percentage of age, over 65 years old in this group was the highest (39.4 percent) and also included the highest percentage of men (40.8%). It is also characteristic that 86.6% of the informal caregivers in this group were married or living in a partnership.

The cluster is characterized by low participation of family caregivers in support activities. The majority (67.1%) of respondents devoted less than ten hours per week to family caregiving. About half of the respondents (52.7%) had been involved in caregiving for less than three years. With regard to the assessment of their own financial situation, the cluster had the highest percentages for the statements “I am well provided for and can afford quite a bit” (52.6%) and “I do not have to limit myself in any way” (6.6%). With regard to the experience of stress or well-being, the respondents showed a low level of stress (Figure 2). In all three questions, which according to the BSFC are formulated in terms of a positive sense of well-being, the highest levels of agreement were found in this group. These include “I have enough time for my own interests and needs” (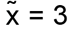(mostly true)), “I can be happy from the bottom of my heart” (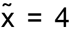(true exactly)) and “I feel I have everything under control” (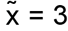(mostly true)).

**Figure 2.**
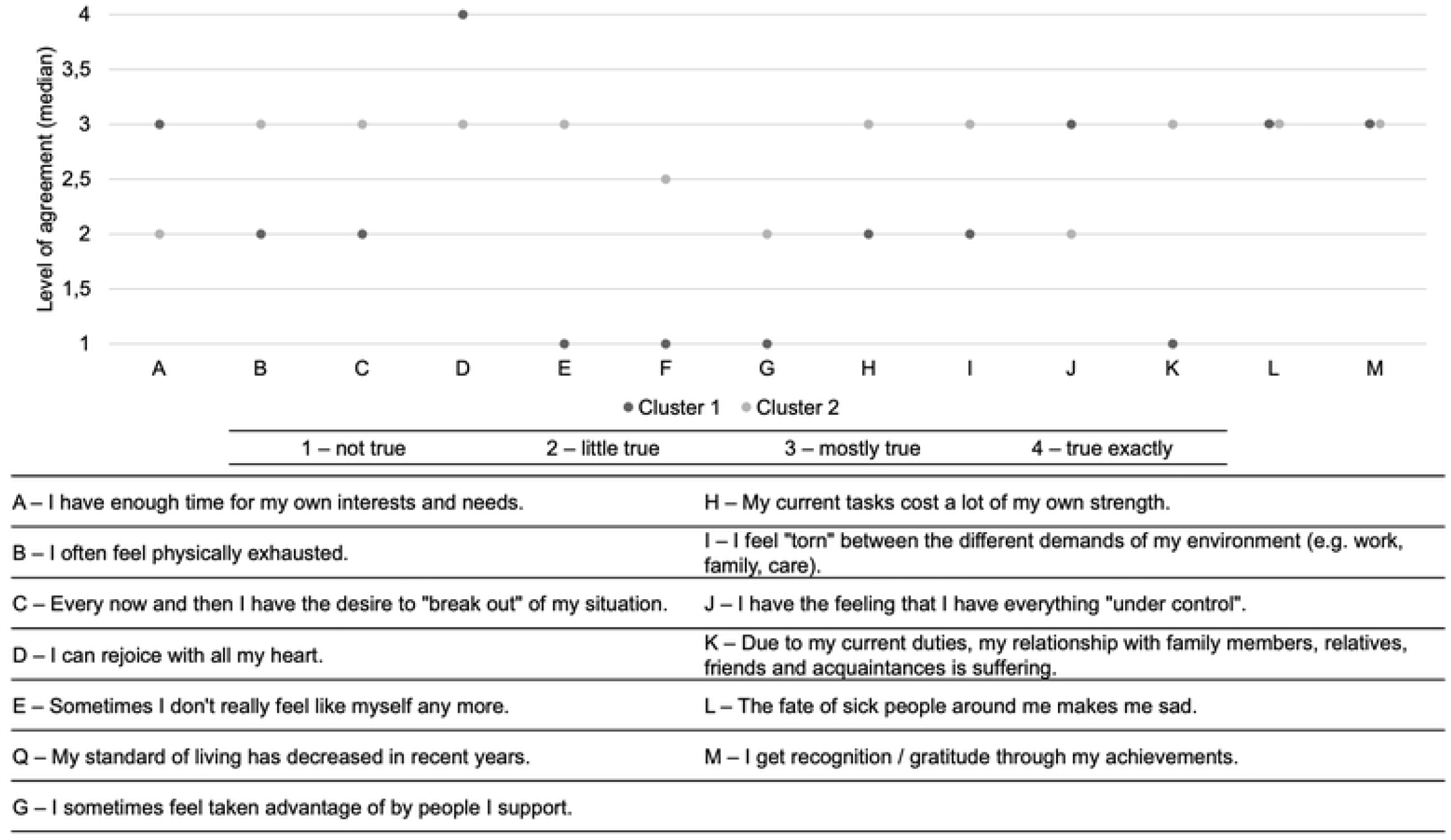
Respondents’ assessment of their personal experience of stress or well-being (n=178)..

At 51.3%, the majority of persons in need of assistance in this group had disabilities in the physical sphere. Relatives in need of assistance mostly included (in-)law mother or (in-)law father (60.5%) as well as other relatives or other close persons (7.9%).

### Cluster 2: intensive assistance, solid to weak financial situation, low well-being

Cluster 2 included 57.3% (n=102) of family caregivers (Table 2). The group included the highest proportion of women (67.6%). The mean age in this cluster was 58.90 years (SD ± 0.884). Again, the majority (75.5%) of respondents reported living in a partnership or being married. Compared to cluster 1, the proportion of single (7.8%) or separated or divorced persons (10.8%) is significantly higher.

Cluster 2 is characterized by a high level of participation in support activities by informal caregivers. Of the participants, 42.2 percent indicated a duration of three to less than nine years in relation to the question of how long they have been helping the person being cared for. Another 21.6 percent indicated a duration of nine years or more. With regard to the weekly care effort expended in the process, compared to cluster 1, the respondents in this group consistently showed the highest values from ten hours per week.

Here, 11.8 percent of respondents stated that they invested 40 hours or more per week in support activities for the person in need of assistance.

Considering their financial situation, the majority of respondents stated “By and large, I get by” (58.8%). A further 27.5% answered with “I just about get by”. With regard to the assessment of the personal experience of stress or well-being in the past four weeks, the respondents showed an increased level of stress (Figure 2). The respondents belonging to cluster 2 always showed the highest agreement values for items that are formulated in terms of a high stress experience.

Only the two statements “The situation of sick people in my environment makes me sad” and “My achievements give me recognition/gratitude” did not differ in the two clusters. Both groups indicated a median agreement level of “mostly true”.

Also in cluster 2, the majority (55.9%) of respondents supported the (in-law) mother or (in-law) father. When asked in which areas the person to be supported had limitations, 45.1% indicated physical limitations and another 47.1% indicated both physical and cognitive/mental impairments.

## Discussion

The aim of this study was to identify and characterize the different views on well-being when taking over nursing services from the perspective of family caregivers. Based on the results of the cluster analysis, two groups of caregiving relatives in Saxony could be identified. The increasing duration and time spent on caring for relatives and the negative trend in the financial situation of informal caregivers are accompanied by an increased experience of stress and a rather low sense of well-being.

In line with the publication by Oltmanns et al. 2016, a positive assessment of one’s own financial situation has a positive effect on subjective well-being [16]. Reconciling care and work often involves conflicts and difficulties. Worrying about one’s own liquid situation can put additional strain on employed informal caregivers, especially in a full-time employment relationship [11]. According to Bohnet-Joschko’s findings, gainful employment can mitigate the negative effects of caregiving and should be maintained if possible. In addition, gainful employment is often reduced due to caregiving in order to be able to provide more extensive care for the relative [17]. It is therefore advisable to further support working relatives in organizing care services and to develop relief programs.

Gratitude and empathy are of great importance in informal care and in care in general. There is consistent evidence in the literature of a positive relationship between self-compassion, as a personal resource, and subjective well-being [18]. If the state of health makes relatives increasingly sad, this can reduce the well-being of informal caregivers. On the other hand, a high level of recognition and gratitude, which informal caregivers receive for their services from the persons in need of help, contributes to an increased creative power and conditions an increased sense of well-being [18].

As the person in need of care becomes more restricted, the caregiving activities become more complex, more extensive and require more time. As a result, the emotional and psychological burden on the family caregiver increases in addition to the physical burden, and they more often feel overwhelmed (cluster 2) [7]. The described physical and psychological burden can be less and less coped with as the age of the family caregiver increases. With retirement age, the proportion of those who support others decreases [17]. However, at older ages (80 years and older), the proportion of caregiving tasks to support provided increases. If people of this age provide help and support to others, it is largely in the form of caregiving activities. Associated with this, the amount of time spent providing assistance also increases with age [19]. The results of the present study reflect this, among other things. While the respondents in cluster 1, who were less involved in caring for their relative, had more than twice the proportion of people over 65, the proportions in the age groups below this in cluster 2 were always the highest.

In accordance with the Bohnet-Joschko cluster analysis, the research results show that informal caregivers who are heavily involved in the care of the relative tend to experience a high level of stress (cluster 2) [11]. A similar picture emerges with regard to the areas of restriction of the person in need of care. Accordingly, relatives who provide support services such as personal hygiene, nutrition and mobility for persons with physical limitations tend to experience high physical stress. High physical strain can lead to physical health problems among informal caregivers themselves [7]. Thus, behind the health effects of home care is the danger that family caregivers themselves become patients. They are also referred to as hidden patients or second victims [3]. In this context, respondents in cluster 1 showed the highest proportion of support in the physical area. The respondents in cluster 2, on the other hand, recorded the highest proportion in the assumption of care activities, with both physical and mental limitations. When caring for relatives with mental limitations, the informal caregiver may experience increased stress and emotional pressure due to social isolation and constant availability [11]. Despite the differences, the two clusters are relatively homogeneous with respect to the domains of limitations. Preventive measures, such as the early teaching of knowledge about the activities as well as back- and joint-friendly working methods when taking over basic care services, can prevent potential damage to the health of informal caregivers. For family caregivers, who are particularly exposed to emotional and psychological pressure, information materials and practical aids can offer the opportunity to have more time for themselves and to engage in compensatory activities [11].

When indicating the relationship of the person to be cared for, with regard to the answer option “(mother-in-law) / (father-in-law)”, the proportion was over half in both clusters. Due to the proximity to the person in need of help, the care situation is often emotionally stressful for the adult children [7]. Self-help groups or networks of informal caregivers with similar fates can provide effective support in this regard, offering mutual exchange and support.

### Limitation

With regard to the data collection date in 2019, it should be noted that this was before the Covid-19 pandemic. In addition, there is the general increase in prices as a result of inflation. Due to these multiple pressures, psychological as well as monetary, cluster solutions may have shifted somewhat. Future research could look more closely at and incorporate such a potential difference based on this foundational work.

In accordance with the objective and research question of this study, a selection of variables was made for the cluster analysis. One selection criterion was to exclude variables from further analysis that contained more than 25 percent missing values. If questions were left unanswered by the subjects, all of the subject’s answers were omitted for the cluster algorithm and he or she was automatically excluded from the clustering. The 25 percent criterion was intended to ensure that the sample size for clustering was not disproportionately reduced due to a variable with numerous missing values, which could have caused further bias in the study results. In addition to variable selection, only informal caregivers were selected for cluster analysis. 82.0 percent of the original sample was thus excluded from further consideration against this background.

The silhouette coefficient in the present study was 0.3, indicating mediocre model quality, according to Kaufman and Rousseeuw [20]. A mediocre classification corresponds to a weak indication of cluster structure, according to the study by Kaufman and Rousseeuw, on which the classification in SPSS is based [20]. However, in order to pursue the objective and research question, it was important to include the listed variables in the cluster analysis.

Cluster analyses allow for the basic desire of people to group objects, facts, etc. into homogeneous groups in order to bring order into a previously confusing situation. It should be noted here that the various generalizing cluster solutions should, however, only be used in an advisory capacity. Information and services should continue to be tailored to the most individual situation possible for family caregivers.

## Conclusion

The research results show that informal caregivers in Saxony are not a homogeneous group. They are confronted with different challenges in caring for relatives, depending on their age, financial situation, care effort and situation. The differentiated level of challenges in the activities, the scope and the individual background of the family caregiver are directly reflected in their well-being and stress experience.

In order to achieve targeted burden reduction for family caregivers, cooperation and constructive collaboration between political decision-makers at the macro level, care and health insurance funds at the meso level and, last but not least, the various actors in health and care provision at the micro level is required. Only if these levels are interlinked and typologies, such as the presented research results from, for example, nursing and health science, are used as an empirical knowledge base, can targeted measures to reduce the burden on informal carers be pursued further in the future.

## Data Availability

Dataset is available at: Tim Tischendorf. (2023). Dataset - Characterization of groups of informal carers. A cluster analysis. [Data set]. Zenodo. https://doi.org/10.5281/zenodo.8255875

